# School education during the SARS-CoV-2 pandemic - Which concept is safe, feasible and environmentally sound?

**DOI:** 10.1101/2020.10.12.20211219

**Authors:** Christian J. Kähler, Thomas Fuchs, Benedikt Mutsch, Rainer Hain

## Abstract

The future belongs to children and they need education to shape the future with foresight and intention. Children therefore have the right to education, according to Article 29 of the UN Convention on the Rights of the Child [1]. However, professional education is not everything, because children must also experience their strengths and weaknesses together and educate each other to be responsible and considerate people, so that they become socially valuable personalities. Only in this way can they shape the future in a peaceful and humane way. Therefore, attending school is essential. However, children also have the right to protection and care by their parents and the state, because the welfare of the child must also be given priority in accordance with Article 3 of the UN Convention on the Rights of the Child. The question is therefore how schooling in community schools can be realized during the SARS-CoV-2 pandemic without exposing children to an unnecessary risk of infection. It is not only about the children, because if the children are at risk, then so are their parents and grandparents and ultimately society as a whole. There are numerous concepts that promise safety in schools during the pandemic. When selecting concepts, the costs must of course be weighed against the benefits. People rightly expect an efficient use of resources. This means that either the set goal is achieved with the least possible resources or that the available resources are used to achieve the greatest possible approximation to the goal. In addition to the financial resources, however, the long-term consequences for the state, the economy, the population and the environment under the pressure of the pandemic must also be taken into account. Social cohesion and democracy must not be jeopardized either. Various protection concepts are currently under discussion. Often the advantages are overstated and the disadvantages concealed. Furthermore, some arguments are based on assumptions that are not true. The aim of this study is to provide a comparative assessment of the main protection concepts and to demonstrate, with the help of experimental analyses, the extent to which the protection concepts are effective. We will show that a comparatively high level of safety against infection in classrooms can be technically ensured without exposing children to masks. At the same time, the protection concept makes economic sense and the burden on the environment is comparatively low, so that infection prevention and climate protection do not have to be weighed against each other, because infection prevention and climate protection are political and social goals that have to be achieved together.

## 1. Introduction

Not all schools are the same, which is due to the different ages of the buildings and their technical equipment. There is therefore no uniform protection concept during the SARS-CoV-2 pandemic that can be recommended and implemented equally for all schools. It is therefore necessary to establish a concept under the respective boundary conditions of the schools that promises the greatest possible safety from infection and takes into account the economic framework conditions and the ecological and social resilience. The fact that the ecological aspect is important does not require any explanation [2, 3]. The economic aspect is important because it would not be responsible if current political decisions with long-term burdens were to excessively restrict our children’s scope for action in shaping the future. The expected burdens on children from national debt, pension burdens, EU rescue parachutes, etc. are already considerable, especially in view of the ageing society and the imminent retirement of the baby boomers [4, 5]. Furthermore, a wasteful use of taxpayers’ money always ensures that socially unprofitable investments are made. This not only leads to unnecessary burdens that restrict the scope for action in the future and create dependencies on financial institutions, but also calls into question the legitimacy of the investment. It is therefore essential that a political goal is either achieved with the smallest possible budget or that the goal is achieved as closely as possible if not enough funds are available. To protect the population during the pandemic, three fundamentally different options are currently being considered and the allocation of funds is currently being negotiated in the social and political process.

One possibility is to use the funds for treatment methods to help the sick people recover. Due to the severity and duration of the infectious disease Covid-19, treatment costs are often high and painful for patients [6]. Since this possibility cannot influence the course of infection, there is a risk that the number of infections will increase rapidly during the cold season and put pressure on the medical system, so that optimal medical treatment can no longer be guaranteed. In addition, many patients with other diseases are also affected by rising infection rates if their treatment is postponed and their care suffers under the burden of the pandemic. Relying on Covid-19 treatment alone would only make sense if an effective and well-tolerated drug is available for all patients. However, this is currently not the case and therefore this option is currently not effective. Nevertheless, the development of a drug is of course very important to enable a mild course of infection and healing without suffering and at low cost in the future.

Secondly, there is the possibility of using a vaccine to reduce the severity of the disease in the event of infection. This approach has proven successful in many infectious diseases and therefore great hopes are placed in the development of a vaccine. The promises of the “redemptive” vaccine are sometimes taking on religious characteristics in the media. However, it must be emphasized that this concept is associated with two major risks. On the one hand, the population must also want a vaccination and inject it practically. The extent to which the population is willing to do so cannot be predicted. On the other hand, a highly effective and well-tolerated vaccine must first be developed, tested, produced and made available worldwide. Currently, however, there is no vaccine that provides effective protection against SARS-CoV-2 infection. The development and testing of a vaccine is usually a long process, so this route may seem promising, but in the current situation it cannot contribute to the management of the pandemic. It would therefore be fatal to hope for a vaccine alone to counter the pandemic. It is also impossible to predict whether a vaccine will provide comprehensive protection at all or only protect to a certain percentage. Even if the current expectations of a vaccine are exaggerated, the development of a vaccine is of course very important because even if not all people could be vaccinated and/or only some of them would lead to immunity, a vaccination would help to relieve the medical system and possibly delay the spread of the virus.

Thirdly, an attempt can be made to prevent the transmission of the virus. If the infection rates are sufficiently low over a longer period of time, there is a basic hope that SARS-CoV-2 will become meaningless. However, this hope also resembles a religion of redemption, because the social efforts necessary for this are not made in a free society. Nevertheless, in the current situation it is very important to minimize the number of infections in order to prevent human suffering and reduce the burden on the health care system and the social, economic and public spheres [6]. But how can this be implemented in practice?

One way is to isolate all infected persons at a time until no one is infectious any more. Even if a rapid test were globally available that could immediately and with 100% reliability prove whether a person is quarantined or not, this way would be practically impossible. Nevertheless, a reliable rapid test would of course be very useful, as it could help to slow down the infection process and prevent unnecessary quarantine periods. Therefore, the investment to develop a rapid detection method would also make sense.

Another way is to effectively interrupt the infection pathways so that infection can no longer take place. This approach requires that the infection pathways are known and that techniques exist that are able to effectively prevent SARS-CoV-2 transmission. Furthermore, people must be willing to use the technologies. Without public acceptance, all concepts are doomed to failure. Even if this approach cannot lead to the complete disappearance of SARS-CoV-2, it is currently the most effective instrument to contain the incidence of infection and thus the costs for the state, the economy and society. This is important because infection prevention allows time to be gained in developing a drug or an effective, safe and well-tolerated vaccine. Since it is impossible to predict how long it will take to develop a vaccine or drug, it would be negligent to financially marginalize the only currently effective protection option, because these technologies and their use alone determine the course of infection in the coming months and perhaps years. Failure in this area directly leads to high costs and suffering in other areas.

According to the current state of research, SARS-CoV-2 is mainly transmitted via droplets and aerosol particles that are produced when breathing, speaking, singing, coughing or sneezing and are inhaled and exhaled through the air we breathe [7, 8, 9, 10]. In order to establish effective measures to prevent an infection, it is initially useful to distinguish between a direct and an indirect infection. Direct infection can occur when many emitted droplets and aerosol particles containing infectious viruses are inhaled over a short distance (less than 1.5 m) by an uninfected person. The smaller the distance, the greater the viral load and thus the risk of infection. This is simply a consequence of fluid mechanics, because with increasing distance from the mouth, the exhaled air jet expands due to friction and entrainment effects, slows down due to the conservation of momentum and the viral load is additionally reduced by superimposed turbulent mixing processes and diffusion. Therefore, distances offer a very good protection against this transmission path. In addition, the duration of the load also plays an important role, because inhaling a high virus concentration for a short time is just as dangerous as inhaling a low virus concentration over a longer period of time. Ultimately, the product of virus concentration × time is the decisive factor. Whether or not an infection occurs depends on the minimum infection dose, i.e. the number of viruses required to actually cause an infection [11, 12]. Direct infection can occur both indoors and outdoors. However, direct infections are very rare in outdoor areas, as convective and thermal air movements impede the straight-line virus propagation to the opposite side and ensure rapid removal of the viruses.

Indirect infection can only occur if the viral load in the room multiplied by the time the person remains in the room exceeds the infectious dose [11, 12]. Consequently, this route of infection can only occur indoors if the room volume is small in relation to the number of infected persons. In large factory or warehouse buildings or huge churches, even many infected persons will not be able to produce an infectious virus dose in the room and therefore the indirect risk of infection in these rooms is insignificant. It is clear that an indirect infection cannot be prevented by keeping a safe distance to infected persons, since the air in the room is contaminated with infectious viruses everywhere. Under these conditions, protection against infection can only be achieved by short residence times or technical aids.

In addition to direct and indirect infection via aerosol particles, there is also the infection path via contact. However, this path of infection is already efficiently prevented by the changed social behavior during encounters (avoidance of shaking hands and other contact) and by general hygiene measures, so that it does not need to be considered further here. If we consider only the direct and indirect path of infection, the question arises as to which concepts can effectively prevent these two infection paths under the conditions of school instruction.

## 2. How can the risk of infection in classrooms be reduced?

Answering the question first requires an understanding of the essential protection concepts and their advantages and disadvantages. In the evaluation we consider in particular the safety against infection, the costs of the measure, the technical and social feasibility and the environmental compatibility.

### Protection concept I: Free ventilation, air conditioning system and CO_2_ indicator

The simplest concept is to carry out the school lessons without any additional protective measures and to reduce the possible virus load in the room only by regular free ventilation via open windows. This variant initially appears to be very cost-effective and easy to implement, since classrooms are usually equipped with windows. It is therefore not surprising that this concept is often presented in a very positive light. The practical implementation of this concept seems to prove its advocates right, as the infection rates are not currently rising sharply. However, this is not because this concept offers security against infection, but because the current infection figures in many countries are quite low and infections are therefore quite unlikely. If the infection figures increase in autumn and winter, the weaknesses and risks of the concept will immediately become apparent.

The biggest shortcoming of the protection concept is that no precautions are taken to prevent direct infections. Neither an increase in the distance between students, nor breathing masks, mouth and nose covers or face shields are used. Furthermore, the possibility of reducing the viral load in the room by regular free ventilation is also overestimated. Free ventilation is only physically effective if there is either a large temperature difference between indoors and outdoors or the wind blows in front of the windows [13, 14]. A temperature difference often does not exist and if it exists, it is quickly reduced by free ventilation, so this mechanism is usually only effective for a short time. The air exchange will therefore take a correspondingly long time, as we will prove experimentally in chapter 3. The wind in front of the window is also rarely strong enough to ensure adequate ventilation. Since the effectiveness of free ventilation depends on factors that cannot be influenced (temperature, wind, size/position of windows), the question remains as to how to ventilate when these physical mechanisms cannot be used.

Since ventilation often does not work effectively, it is recommended in the warm season to open all windows very wide during the entire time the room is used. Cross-ventilation is particularly effective [15], but windows on opposite sides of the room are rare. If, in the warm season, even with wide open windows, no efficient ventilation success can be achieved via physical mechanisms, it is recommended to place a fan in a window to supply the room with virus-free outside air. The removal of the viruses from the room will then automatically take place through the other windows. The extraction of the room air with a fan is less recommended, even if it seems physically reasonable at first, at least if the indoor air is warmer than the outdoor air. The reason for this is that the slight negative pressure in the room during the extraction process causes room air to be sucked in from other parts of the building through door gaps and ventilation ducts etc. Since this air can be contaminated in the adjacent rooms, it is recommended for safety reasons that the pressure in the room be increased by blowing in outside air [15].

During the warm seasons, free ventilation is certainly the most cost-effective method to limit the increase of the virus load in the room and to prevent indirect infection. During the cold season, however, this ventilation concept leads to colds and the well-being of people is impaired. Continuous ventilation is therefore not an option in autumn and winter. If you switch to regular push ventilation, you must always remember to do so and the students must be willing and able to do so (in many schools the windows cannot be opened). In addition, the question of how often and for how long the school should be ventilated arises.

Regularly a CO_2_ indicator is advertised as a solution to the problem. It is assumed that the CO_2_ value correlates with the virus load in the room. But this assumption is wrong. First of all, it should be remembered that the viral load depends on the number of infected persons in the room, their length of stay and their activity. If, for whatever reason, it is assumed that the viral load in a room is reached after t minutes if only one person is infected, then in the case of two infected persons the ventilation would have to be started after t/2, even though the assumed critical CO_2_ value is only reached after t minutes. If there are more infected persons in the room, then the time is further reduced according to the number N of infected persons according to t/N. This does not even take into account the activity of the persons. Therefore, a CO_2_ display is only a rough measure of the virus load if the number of infected persons in the room is known. But this number is unknown and therefore the CO_2_ measure is no indicator for the risk of infection. The problem can also be illustrated with another example. If there are 10 healthy people in a room, after a certain time a certain CO_2_ concentration will be reached and the viral load is zero. If 5 persons step out and 5 infected persons step in, the CO_2_ concentration will hardly change, but the viral load in the room will increase very quickly and with it the indirect risk of infection. The desire to offer a solution for 25 Euro per classroom, which suggests safety, is understandable, but this supposed solution does not serve the purpose. It could be argued that the number of infected persons is small and therefore it could be expected that in most classes there is no or at most one infected person statistically. This is certainly true at present, but two things must be taken into account. Firstly, it is to be feared that the currently low number of infected persons will rise sharply in winter. This will also increase the probability that infected persons will be in the classrooms. Secondly, it must be taken into account that an infected child is very likely to infect other children if there are no adequate protective measures in place. This can be explained by the fact that the children are very familiar and get close to each other in class over a long period of time. It is therefore to be feared that especially in schools with insufficient protection superspreader events will become very important [16].

However, the CO_2_ indication is extremely useful to determine whether ventilation is successful by shock ventilation and thus the necessary ventilation time can be determined. If the CO_2_ value does not change significantly during shock ventilation, it is clear that free ventilation does not work because either the temperature inside and outside is the same or there is no wind. What to do in this case is unfortunately not answered by the advocates of the concept.

The main argument against free ventilation during the cold season is the waste of thermal energy. In order to conserve resources and limit global warming, houses are elaborately and cost-intensively insulated and highly efficient heating systems are installed. It makes neither ecological nor economic sense to first implement these measures and then let the thermal energy out of the open window. The demand that the climate targets should be of secondary importance during the pandemic is incomprehensible. Surely the goal must be to find a solution that reconciles infection control with climate protection.

In new modern schools, some of the disadvantages of free ventilation can be counteracted by so-called HVAC (Heating, Ventilation and Air Conditioning) systems [17]. If the systems are operated with 100% outside air and the air exchange rate per hour is more than six times the volume of the room, then the ventilation success is regulated independently of the temperature and weather situation and nobody has to think about regular ventilation and put it into practice. In this respect, these systems offer a significant improvement over free ventilation. Therefore, they should be used and operated as described above, if they are available, in order to create a high degree of security against indirect infection [18]. However, it is clear that these air handling units with 100% outside air and an air exchange rate of 6 per hour do not offer any energetic advantage over free ventilation. It should also be noted that HVAC systems are often equipped with frost monitors that ensure that the systems do not start up in winter when the fresh air rate is 100%. Otherwise they could freeze and suffer damage. In order to solve the technical problem, the HVAC can be operated at low temperatures with a large proportion of circulating air. In this operating mode, they work energetically favourably, but then they become virus spinners, as the viruses are usually not effectively separated, but are passed from room to room. This problem has been known for a long time and needs no further explanation [19, 20, 21, 22, 23, 24]. Recirculation must therefore be avoided or filters must be used that are capable of reliably separating the viruses [17]. It is often argued that filters of class F7 or F9 typically installed in air handling systems are also capable of separating aerosol particles. This is true, but the separation efficiency is not sufficient. According to the decision of the “Laboratory Technology” project group of the Committee for Biological Agents (ABAS), only class H14 filters are suitable for separating viruses [25]. It would therefore make sense to integrate class H14 filters into the air conditioning systems of modern schools, so that safe and energetically favorable recirculation air operation is possible during the cold season. Unfortunately, existing air handling systems cannot be converted so easily, as the volume flow decreases or the systems suffer damage. Whether UV-C radiation could offer a safe solution of the problem, must still prove itself with the large volume flows.

It is also occasionally argued that air exchange rates of 6 per hour are not necessary and at air exchange rates of 1 – 2 the HVAC would also work in winter with a high proportion of outside air. This view must be expressly contradicted. Air exchange rates of 1 – 2 are certainly sufficient to limit the accumulation of CO_2_ in the room and thus enable the students to work in a concentrated manner. However, they are by far not sufficient to provide protection against a dangerous virus [26]. It is not without reason that rooms in hospitals with infectious patients are operated with air exchange rates between 12 and 15 [26, 27, 28]. According to DIN 1946-4, a minimum air volume flow of 40 + 100 m^3^/(h × person) per patient is required in a patient room. Against this background, to assume that children are safe from indirect infection at air exchange rates of 1 – 2 is incomprehensible and almost negligent with regard to the lethal risk associated with Covid-19 disease. The fact is that accepting infections leads to high costs for the healthcare system. The air exchange rate we recommend, which is at least 6 times the room volume per hour, represents in our opinion a good compromise between safety and technical feasibility. If this value should prove to be insufficient, for example because too many people are infected, then it must be raised further.

Some people argue that the air exchange rate is not a suitable measure to minimize the risk of infection, although this number has been used for decades. These people believe that the air exchange rate must be related to the number of infected people in the room. The problem with this approach is that the number of infected people is not known. If you know who is infected, you would send the infected people directly home and not bother with ventilation issues. The advocates of this view obviously do not consider that in reality it is not known how many people are infected in a room. Because this information is not known, it is precisely the air exchange rate that is taken, since it is the only meaningful variable that would allow a high level of safety to be ensured, regardless of the number of infected persons in the room. The argumentation of the people who criticize the air exchange rate is also completely wrong for another reason. Let us assume that the virus load in a room is high because there were just a few infected persons in the room. This means that the indirect risk of infection is correspondingly high when you are in that room. Now the infected persons leave the room. According to the argumentation that only the number of infected persons in the room is important, it would now be safe to enter in the room, because the infected persons are no longer in the room. But that is of course complete nonsense. This example shows that it is not the infected persons in the room that are essential but the virus load in the room. This depends on the number, activity and duration of the infected person in a room. One infected person who stays in a room for one hour can release as many viruses as two people who are in the room for half an hour. These examples show that the reasoning based on the infected persons is completely pointless. It is important to minimize the virus load in the room and for this the air exchange rate is the decisive factor.

In summary, it can be said that a protection concept that relies solely on free ventilation offers only a minimum of security against infection. The concept offers no protection whatsoever against direct infection, although this is the most likely. This is the greatest weakness of the concept. Free ventilation through permanently open windows or with HVAC systems that supply 100% outside air at an air exchange rate of 6 per hour is useful during the warm season to minimize at least the indirect risk of infection. In winter, however, free ventilation is energetically unfavorable. Furthermore, it is doubtful whether there is any willingness at all to ventilate regularly. Furthermore, it is completely unclear how regular ventilation is necessary, since the number of infected persons is not known. HVACs do not offer any energetic advantage in winter if they are operated with 100% outside air and six times the air exchange rate per hour. If they run in recirculation mode, a lot of energy can be saved, but in this mode the systems become virus spinners and endanger the safety of children. This can only be prevented by integrating a class H14 filter, which separates 99.995% of the aerosol particles and thus the viruses when the air flows through once. Alternatively, other technologies can be used (UV-C, ionization, …), but it must be ensured that these technologies are able to inactivate 99.995% of the viruses in a single pass through the device. If the enormous waste of energy of the concept for the prevention of indirect infections in the cold season is taken into account and the fact that each direct infection, which is cheaply accepted here, is connected with substantial costs, then it is not comprehensible, why this uncertain, unpleasant as well as economically and ecologically precarious concept receives so much support in the current discussion.

### Protection concept II: Safety distances

One suggestion that is made time and again is to increase the distance between children in the classroom to reduce the risk of infection. There is no doubt that direct respiratory infection over short distances can lead to SARS-CoV-2 infection. It has also been proven that the direct probability of infection is reduced with distance and is virtually non-existent at large distances (greater than 3 m). Large safety distances therefore offer a very effective foreign and self-protection against direct infection.

A serious disadvantage of the protection concept, however, is that it is practically impossible to realize sufficient safety distances in the classrooms, since the corresponding rooms are not available. Doubling the distances between children and adolescents in both directions would require a quadrupling of the required classroom space. The lack of classrooms could theoretically be solved by running the school in four shifts, but there is a lack of teachers, and their training and employment would involve immense costs.

Another major drawback of the concept is that distances alone in a closed room cannot guarantee safety from SARS-CoV-2 infection. Since the viral load in a room depends on the number of infected persons and their length of stay and activity, additional measures must be taken to limit the viral load in the room air, otherwise indirect infections may occur.

The indirect risk of infection can be counteracted by free ventilation or air conditioning systems in the warm season, as already mentioned, as all windows can be opened wide or the air conditioning system can be operated with 100% outside air and an air exchange rate of at least 6 per hour. The problems during the cold season have already been discussed in detail.

It can therefore be concluded that the concept of safety distances is neither safe nor practical to implement and certainly not financially viable. The necessity of free ventilation also makes the concept appear disadvantageous from an economic and ecological perspective during the cold season. The only way to realize this concept is to provide home schooling for the students. However, this approach is not recommended for several reasons. On the one hand, the school not only provides education, but also makes a very large contribution to making the children socially acceptable. Online schooling or lessons by the parents cannot achieve this. Furthermore, schooling by parents is highly inefficient. On the one hand, they are usually not educators who are really qualified to teach material. Furthermore, not all parents have the necessary expertise. This would lead to a great educational injustice. It must also be taken into account, however, that many parents are already working to capacity due to their jobs or family activities. To deduct this valuable workload now in order to teach the children in individual lessons is highly inefficient and therefore economically disadvantageous, not only because of the necessary familiarization with the curriculum and the poorer teaching of the material. If you also consider that one teacher teaches 20 – 30 children in a school, then it is certainly not sensible to move to a system where the ratio is 1 : 1. Although a domestic protection concept can offer a high degree of safety from SARS-CoV-2 infection, the disadvantages for the children, parents, the economy and society are far too great.

### Protection concept III: Use of FFP2/3 masks in class

Direct and indirect infection can be effectively prevented with the aid of high-quality particle-filtering respirators (FFP2/3 or better), as these respirators reliably trap droplets and aerosol particles during inhalation and exhalation up to a defined size class, provided they are firmly and tightly fitted to the face [29, 30]. If these masks are used without an outlet valve, large safety distances between persons are not required to prevent direct infection. In addition, nothing needs to be done to prevent indirect infections caused by an increased viral load in the room, since particle-filtering respiratory masks also provide reliable protection against this transmission route [29, 30].

The protective function of these masks is sometimes doubted because they are not 100% leak-proof. It is a fact that particle-filtering respiratory masks are part of occupational safety in hospitals, laboratories, isolation wards, operating rooms and many technical work areas where fine dust and hazardous substances are handled (e.g. grinding, welding, soldering). The argument that the SARS-CoV-2 viruses cannot be reliably separated because the viruses are smaller than 0.16 μm is not correct either, because SARS-CoV-2 is transmitted via droplets or droplet nuclei and these are significantly larger than individual viruses and can be separated quite reliably by suitable particle-filtering masks [30]. It should also be noted that very small aerosol particles often do not carry viruses and even if they do carry a virus, many of these very small aerosol particles would have to be inhaled to cause infection [31]. It is estimated that a dose of at least 500 – 2000 viruses is required to cause a SARS-CoV-2 infection [11, 12]. If all persons in a classroom wore these masks, there would be double protection. On the one hand, hardly any viruses from an infected person will enter the air in the room. On the other hand, the few viruses that do make it into the room air could hardly overcome the mask barrier of an uninfected person. Therefore, it can be assumed that this protection concept offers a very high degree of security against a SARS-CoV-2 infection.

A major disadvantage of particle-filtering masks without a valve is that they make breathing difficult and wearing them can be uncomfortable in the long term. In order to prevent overuse, the masks should only be worn for a maximum of 3 × 75 minutes per day, with a break of at least 30 minutes every 75 minutes [32]. They are therefore hardly suitable for schools to provide lasting protection.

However, it should also be borne in mind that these masks not only make breathing more difficult and are sometimes uncomfortable, but also cause considerable costs in the long term. If a school class with 25 children uses FFP2/3 respirators daily for a unit price of 4 €, then 200 school days per year would result in a total of 20000 € per class per year or 800 € per child per year. In addition to these costs, it must also be taken into account that the masks produce a lot of waste, the processing of which causes further costs.

It remains to be noted that although FFP2/3 respirators that are tightly and closely fitting to the face provide very good protection against direct and indirect infection, wearing them permanently has a detrimental effect on the health and well-being of the wearer. Therefore the concept is not really feasible. Furthermore, wearing respiratory masks makes neither economic nor ecological sense. However, it must be emphasized that these respirators must be worn as soon as the children leave their place and walk through class or school or go to their place in the morning or after a break! The wearing time is then minimal and the mobile protection against droplet and aerosol particle infection is maximal. As the masks are only worn for a short time, they can be used for 2 – 3 weeks without any problems.

### Protection concept IV: Room air purifier + mouth-nose cover / face visor

Currently, there is an increasing number of concepts being discussed that use room air cleaners/purifiers [33] or disinfection devices [34] to protect against indirect infection and provide individual protection measures to prevent the direct path of infection. Indoor air cleaners with class H14 filters and disinfection devices with electrostatic filtration, UV-C and ionization unit were analyzed in two studies to test their effectiveness [33, 34]. The main result of the study is that the viral load in the room is reduced very quickly and the retention time of the viruses after release is short if three criteria are met:

1. The volume flow of the devices must be at least six times the room volume per hour. For an 80 m^2^ room with a volume of 200 m^3^, the unit must therefore have a capacity of at least 1200 m^3^/h. Low air exchange rates may be sufficient to remove largely harmless contaminations such as CO_2_, pollen or fine dust from the room air, but significantly higher air exchange rates are required for the fast and safe removal of dangerous viruses. An air exchange rate of six times the room volume per hour already represents a compromise between safety and technical feasibility. In hospitals or laboratories where the room air is contaminated with dangerous viruses, air exchange rates of 12 – 15 are usually required [26, 27, 28]. If it turns out that an air exchange rate of 6 per hour should not be sufficient because, for example, the number of infected persons per room is large on average, then higher air exchange rates must be implemented. It should also be remembered that the air exchange rate selected on the device only corresponds to the actual air exchange rate if there is good air mixing in the room.
2. The filter must have a filter performance according to EN 1822-1 at the required volume flow rate in order to separate 99.995% of the aerosol particles from a diameter of 0.1 – 0.3 μm or the viruses must be inactivated with this effectiveness when passing through the device once by means of UV radiation or electrical charges. The droplets produced when breathing, speaking, singing and coughing and the droplet nuclei formed from them by evaporation are only up to a few micrometers in size and can therefore only be separated almost completely and efficiently with really high-quality filters of class H14 [25]. The argument that this separation efficiency can also be achieved with a simple filter by multiple filtering could not be confirmed in earlier experiments [33]. However, this argument is also inappropriate, since this approach would require a doubling or even multiplication of the required air exchange rate in order to achieve a comparable level of safety from infection as that achieved with a class H14 filter in a single flow. This is hardly technically feasible and the comfort in the room would suffer from the noise and drafts.
3. The device must be sufficiently quiet so that it does not disturb during operation. If the noise disturbs, there is a risk that the unit is switched off completely or is not operated with the required air flow. In this case, there is no high degree of safety against indirect SARS-CoV-2 infection. The volume of the devices usually depends on their size. Devices with large fans are quiet and small devices with high fan speeds are loud at the same volume flow.

Small and cheap devices usually do not meet these three requirements. Therefore, most of these devices cannot provide a high level of protection against SARS-CoV-2 infection. Supposed quality seals, which many cheap and compact devices have, such as HEPA filters, suggest safety, but do not offer it, because it depends on the filter class according to the filter standard EN 1822-1. Certificates with cryptic characters should not be trusted and assurances that foreign certifications or other regulations comply with the EN 1822-1 standard should not be believed. Well-known manufacturers may appear trustworthy, but in the end it is the technical data of the device and not the reputation of the manufacturer that counts. It is important to pay attention to the three criteria mentioned above when buying such devices. Only if the three criteria are met, the devices offer a very good protection against indirect SARS-CoV-2 infection in schools and other environments. However, it is not possible to verify whether the information in the data sheets is correct.

A major advantage of the room air purifiers and disinfection devices is that they permanently ensure a low virus load in the room without having to worry about opening windows and without affecting the well-being in the room. In addition, unlike free ventilation with windows, they also ensure that there is a real reduction or inactivation of the virus load, which often cannot be guaranteed by open windows. They also offer the advantage over HVAC systems that are permanently integrated into buildings and operated with little or no outside air, that the viruses are really eliminated or inactivated and are not distributed through ventilation shafts in the building. Occasionally, the accusation is made that the room air cleaners and disinfection devices are “virus spinners”. This is certainly true for simple devices with poor filters. However, devices with class H14 filters cannot be virus spinners, because the filter separates almost all aerosol particles. The assumption that the filter is permeable to viruses or that parts of the filter become detached are wrong assumptions, because the filter would not be a filter if it could not reliably separate the aerosol particles, viruses, bacteria, pollen and fine dust. This is why these filters are also used in hospitals and laboratories. The ABAS resolution 16/2010 of December 2, 2010 clearly states the requirement for the separation of viruses: “The HEPA filters should at least correspond to class H14 according to DIN EN 1822-1. On the basis of the risk assessment, H13 filters can also be considered if there are special reasons, e.g. exclusively bacteriological work” [25].

As a result of the scientific investigations [33, 34], it is clear that room air cleaners and disinfection devices with a volume flow per hour that corresponds to at least six times the room volume and high-quality filters of class H14 represent a very sensible technical solution to greatly reduce the indirect risk of infection by aerosols in classrooms. They can also be used as a support in buildings with low-performance HVAC systems. Even in winter, when the HVAC systems are not able to handle large volume flows with outside air, room air purifiers and suitable disinfection units can be used in addition to ensure the desired safety from indirect infection. When installing the units, it is only necessary to ensure that the intake area of the unit is not blocked and that the ceiling is as level as possible so that good air distribution can be guaranteed [33].

However, it should be noted that room air cleaners and disinfection devices are only suitable for minimizing the indirect risk of infection. Additional measures must therefore be taken to prevent the direct risk of infection. For the prevention of direct infections, mouth-and-nose covers, surgical masks or transparent face visors are suitable. These precautions have the effect of strongly limiting the forward spread of exhaled air [29, 31]. Therefore, they offer good protection against foreign bodies when talking face to face, even if the person coughs. However, it must be remembered that these technical aids do not significantly filter the aerosol particles out of the exhaled air because, on the one hand, the filtering effect of the material is too poor and, on the other hand, because the masks do not seal the face firmly enough or not at all. Since the flow essentially follows the path of least resistance, the air enters and exits primarily through the gaps at the edge of the mask when breathing. In the case of the mouth-and-nose cover, this is to the side and thus to the seat neighbour [30]. For this reason, mouth-and-nose coverings do not offer any significant protection for either the other person or for themselves in the classroom situation. Consequently, this method of protection is not recommended when people sit directly next to each other for long periods of time.

Mouth-and-nose coverings and surgical masks are not only unsafe because of the lateral leakage of the breathing air, but also because they are rarely put on properly. In practice, the nose is often not covered and the gaps on the face are much too large. Furthermore, they are perceived as annoying when worn for a longer period of time, because the material often itches or pinches and is uncomfortable in damp conditions. These irritations and comfort disadvantages cannot only reduce the feeling of well-being, but also the attention to the lessons.

With a face visor the breathing air is primarily redirected downwards and partly upwards. Considering the seating arrangement in the classrooms, this can be considered safer than a lateral escape of the breathing air. In a recent publication, facial visors were evaluated as unsafe because breathing air escaped significantly forward despite the face visor [35]. However, it should be emphasized that the face shields were tilted strongly forward during the experiments and were therefore not put on correctly. The face visors also have the great advantage over mouth-and-nose covers and surgical masks that they are more comfortable to wear and the facial expressions are visible. Nevertheless, face masks are not practical in teaching, because they hinder the tilting of the head. This restricts the freedom of movement in the classroom and especially when writing, the face visor can be a hindrance. Therefore this protection concept can be classified as safer than protection concept I and despite the investment costs for the air purifier of 2000 to 4000 € and the face visors it is also cheaper than the other concepts, because the number of direct infections is reduced, compared to protection concept I, no energy is wasted through windows or air conditioning systems, as in protection concepts I and II and because no expensive masks have to be purchased, as in the case of protection concept III. But a real normality of school operations cannot be established in the long run even with this concept, because the use of face visors restricts and hinders too much.

### Protection concept V: Room air cleaner + transparent protective walls

Little attention has so far been paid to a concept that minimizes indirect infection with a room air purifier [33] or disinfection device [34] and realizes direct infection between neighboring children via transparent partitions that can be easily mounted on the table. The advantages of indoor air cleaners and disinfection devices have already been described in detail. The disadvantages that arise if the three important criteria are not taken into account when purchasing the equipment have also been clearly stated. One question that remains is whether these air purifiers and disinfection devices can also effectively limit the indirect risk of infection when classrooms are filled with tables and chairs, people with backpacks, etc. Only if this is guaranteed can indoor air cleaners and disinfection devices make a meaningful contribution to infection control in schools. Furthermore, the question is how direct infection protection can be realized in the classroom without the described disadvantages of mouth-and-nose covers, surgical masks and face visors.

The most effective protection against direct infection is provided by solid partitions between adjacent persons, as they are completely impermeable to aerosol particles and viruses. If these partitions are large enough, it is also very unlikely that the exhaled air will rise, flow over the boundary and then sink down to the adjacent space. Thus, partitions provide the best possible protection between adjacent people in the classroom. The table neighbours can get as close as they like through the barrier and can also talk and cough face-to-face without getting infected. Furthermore, the partitions do not disturb the students’ working methods, facial expressions are visible and the room is completely visible, provided that the protective walls are made of transparent Plexiglas. If the partition walls are well dimensioned and executed, they do not pose a problem in case of fire, because the possibility of escape is hardly limited.

A disadvantage of these walls is that during direct face-to-face conversations between the table neighbours, the airflow flows along the wall, detaches at the end and then continues to flow towards the next row of seats. This problem can be completely eliminated by placing the tables in a U shape. But what if there is not enough space in the classroom for this arrangement? If the transparent partition wall is provided with a circumferential edge that protrudes about 30 mm on both sides, the air that spreads along the wall is deflected by 90° by this edge and a corner vortex is formed that remains in place until it is sucked in by the room air purifier. This simple protective measure can also reduce the risk of infection for people sitting in adjacent rows of seats. In addition, the distance between the rows of seats should be maximized so that the persons in the front rows are not exposed to an increased risk of infection even when looking towards the teacher.

The protection concept, consisting of a room air purifier plus a protective wall, seems promising, as it promises safety from direct and indirect infection without the children having to wear annoying masks during lessons. Masks are only necessary when the children leave their place and walk through the school. The facial expressions of the children and adolescents would thus be recognizable in class and the room would appear transparent. This concept also means that allergy sufferers will have fewer complaints, as pollen and fine dust are also separated and infection with other infectious diseases is also reduced. The purchase and operating costs are comparatively low and no energy is wasted or waste produced, so that this concept is also advantageous from an economic and ecological perspective. Furthermore, the concept is easy to implement and provides a high level of safety without human intervention. Since there are many producers of such devices in Europe, it can be assumed that the devices are also available in large quantities. Furthermore, the use of these devices would not only ensure safety in classrooms and offices, but also secure jobs in Europe. If the technology proves itself in our schools, it is likely that other countries will follow suit and perhaps even buy the technology in Europe. The positive aspects for the economy and employment are obvious. However, since this concept has been analysed far less than the other protection concepts, a final evaluation must first prove the effectiveness of the protection facilities in a generic classroom situation.

## 4. Test setup and execution of the concentration measurements

In order to qualify the protection concept, consisting of a room air purifier and protective partition wall, for school operation, an 80 m^2^ generic classroom with a room volume of 200 m^3^ and 24 seats with tables was built. The arrangement is shown schematically in Figure 1. Two of the seats are not designed, because the laser measuring system was installed there to determine the concentration of the aerosol particles. Since the decrease of the aerosol particle concentration in the room according to [33, 34] depends only very slightly on the position, only one measuring device was used for the investigations. In addition, the number of particles in the corners of the room was determined with an oprical particle analyser to prove that the particle concentration in the room is homogeneous.

**Figure 1:**
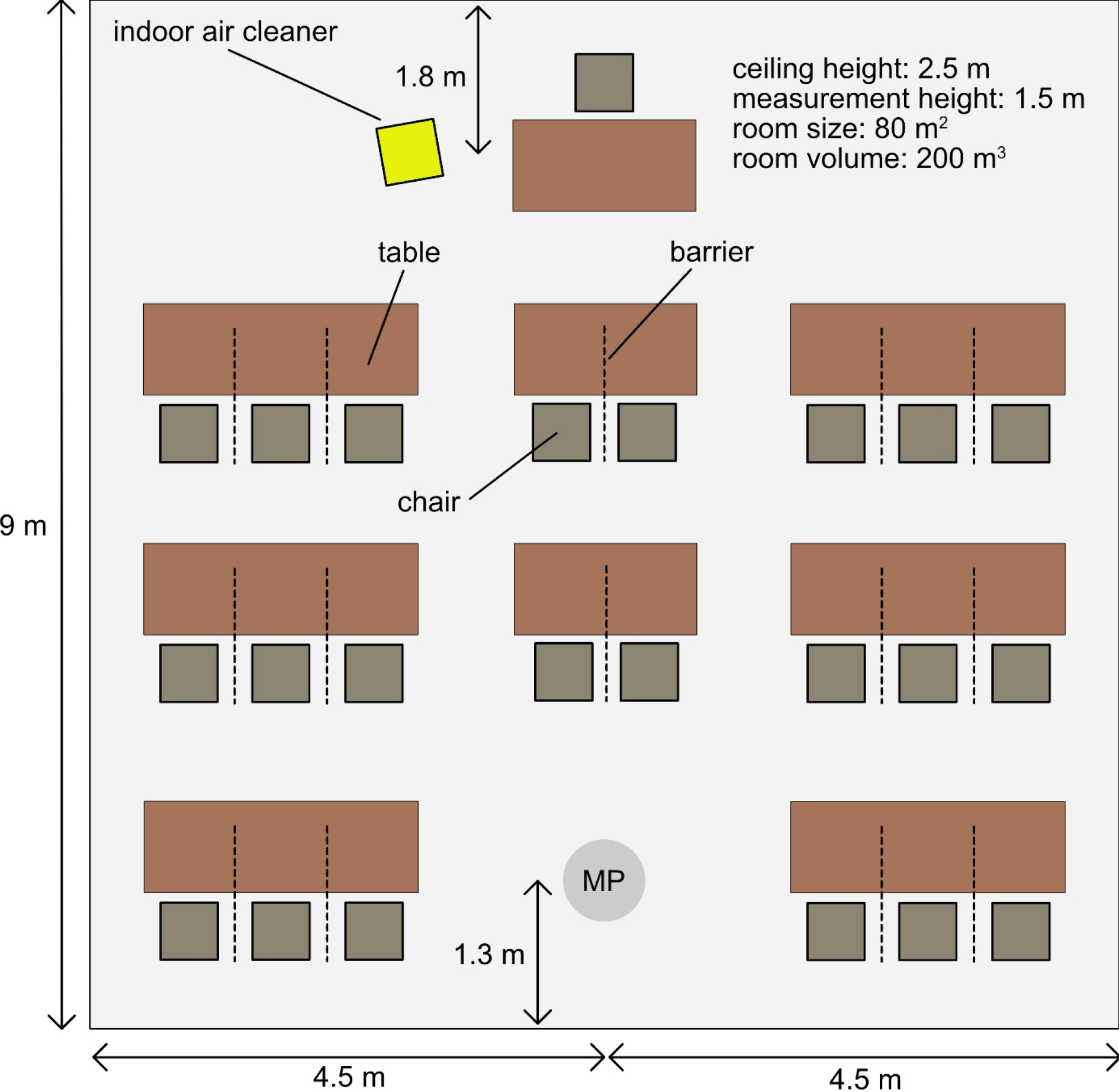
Arrangement of the components in the room for concentration measurements

The room air cleaner (TROTEC TAC V+) was positioned close to the console table and operated with a volume flow of 1200 m^3^/h to achieve a nominal air exchange rate of 6 per hour. The actual air exchange rate can be lower if the room air is very quiet. It is therefore good if people move around in the room and ensure a good mixing. Furthermore, a measurement with the room air purifier switched off was analysed as a reference (0 m^3^/h) and a comparison case with 1000 m^3^/h. In addition, for comparison purposes, it was analysed how quickly the concentration decreases if only a door 2.05 m high and 0.89 m wide is opened to the maximum. This measurement illustrates how effective free ventilation is when the temperature difference between inside and outside is small and there is no wind in front of the door. These conditions occur very often in the temperate seasons. But also in winter, when free ventilation is frequent, this situation is close to reality. In addition to measuring the concentration, the CO_2_ content in the air, the temperature and the relative humidity as a function of time were also measured. During the experiments, with 13 people in the room, the temperature increased by about 1 °C and the CO_2_ concentration from 0.08 % to 1.7 %.

The decay constant, the half-life and the mean residence time of the aerosol particles in the room can be determined from the temporal course of the number of aerosol particles in the measuring volume [13, 14]. The value of the decay constant theoretically corresponds exactly to the air exchange rate. The half-life is a measure of how long it takes for the number of aerosol particles at the measurement position under consideration to decrease by 50%. The mean residence time is the average time the released aerosol particles take to travel from the respective measuring position to the deposition site in the sterilizer. Due to the large distance between the room air purifier and the measuring system, the measured value is close to the maximum value, i.e. the concentrations were measured at the most unfavourable location. The smaller the distance between the persons, the shorter the residence time of the released aerosol particles in the room until they are separated or the viruses are inactivated.

To determine the temporal decrease of aerosol particles in the room air, the room was first homogeneously nebulized with very long-lived di-2-ethylhexyl sebacate (DEHS) aerosol particles with a size distribution between 0.1 – 2 μm and a mean diameter of about 1 μm [36]. The size distribution of the aerosol particles is comparable to the aerosol particles emitted during breathing, speaking and singing [37, 38, 39]. The aerosol particles were illuminated with a Litron Nano L 200-15 PIV Nd:YAG double pulse laser and digitally stored with a PCO.edge 5.5 sCMOS camera with 50 mm Zeiss lens. The camera and the laser were centrally controlled by the software DAVIS from LaVision. The acquisition rate for the measurements was 1 Hz. The number of particle images on the sensor corresponds to the number of aerosol particles in the illuminated measurement volume. If the room air purifier is switched on for the measurement, the result of the measurements is the number of aerosol particles in the measurement volume as a function of time. Figure 2 (left) shows a distorted panoramic image of the experimental setup.

**Figure 2:**
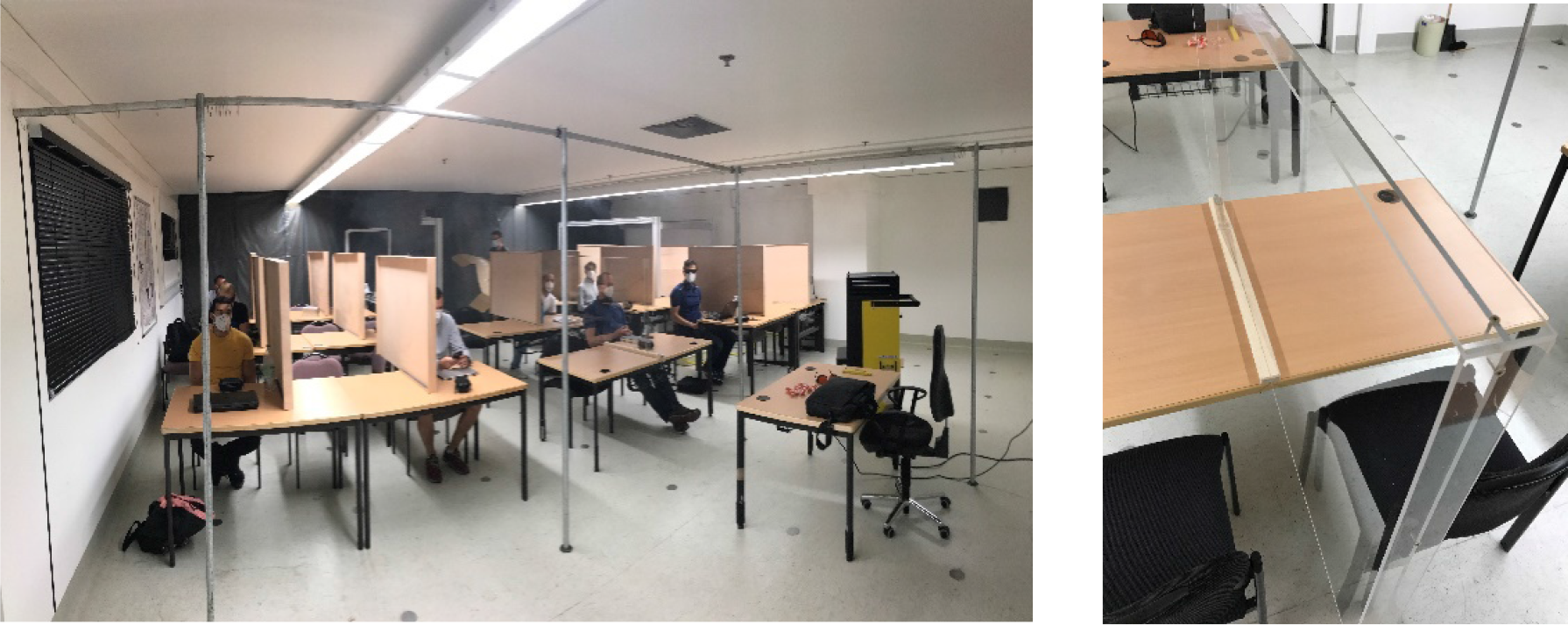
Optically distorted panoramic image of the experimental room (left) and a detailed view of the transparent partition wall with surrounding edge (right). The other partitions were made of wood, as shown in the left picture.

Three different room situations were considered for the analysis.

1. classroom without persons
2. classroom with 13 adults, bags and backpacks
3. classroom with 11 adults, bags and backpacks and partitions between adjacent seats at a table.

The number of persons in the room was limited due to the security regulations of the Bundeswehr University Munich. The partition walls have a size of 1.2 m × 0.8 m. For the investigations, one protective wall was made of Plexiglas and the others of spearwood. The special feature of the protective walls is a circumferential edge that protrudes approx. 30 mm on both sides, see Figure 2 (right). This edge has a very important protective function. If you speak, blow or cough against the protective wall, the air flow is redirected at the wall and then initially spreads tangentially along the wall. If there is no boundary at the edge of the wall, the air simply continues to flow beyond the edge and can reach people in the rows of seats in front or behind it, as long as it flows fast enough. If, on the other hand, the circumferential edge is used, it deflects the air flow and creates a corner vortex at the edge of the partition, which remains there until it has dissipated due to friction effects at the location. The aerosol particles cannot therefore reach the adjacent rows of seats. Furthermore, the dilution of the viral load is promoted by the vortex.

In order to make the partitions easy to set up, they were held in place with a slotted base, which was attached to the table with double-sided adhesive tape and a small screw clamp. The foot was extended to the other edge of the table, so that a small screw clamp could be attached there for fixation to prevent the children from bumping their legs. The protective wall protrudes approx. 0.6 m behind the end of the table so that the protective wall protects the children safely from direct infection even if they lean back or tilt slightly with the chair while talking face-to-face.

### Experiments with homogeneous aerosol particle distribution in space

Figure 3 shows the decrease of the measured concentration as a function of time in normalized representation. The course of the reference measurement (0 m^3^/h) shows that the aerosol concentration in the room hardly decreases if the room air purifier is not operated (black dots). Infectious aerosol particles in the room can thus become a great danger, since the small aerosol particles are hardly deposited on the walls and the floor.

**Figure 3:**
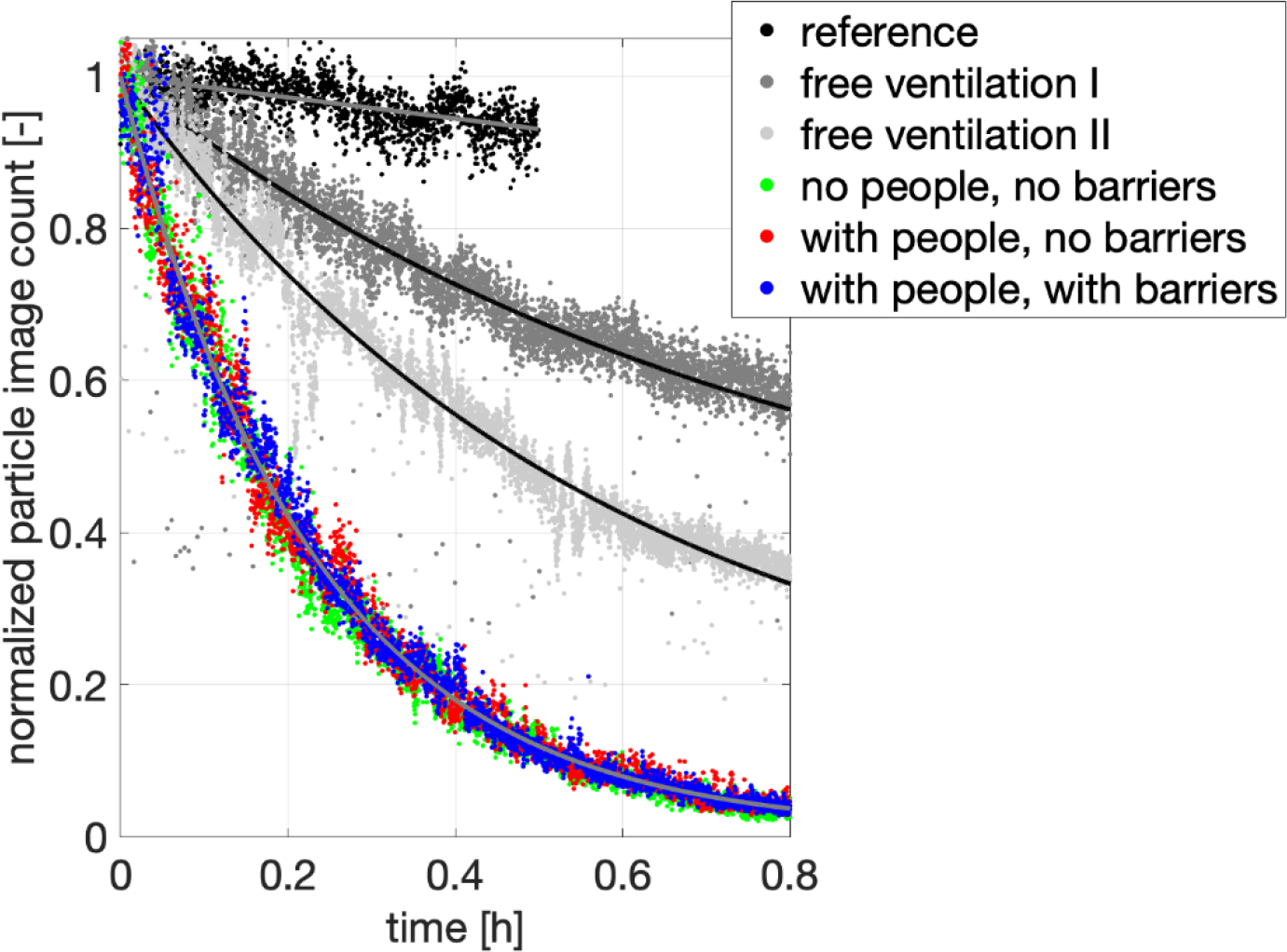
Decrease of aerosol particle concentration over time for different configurations and free ventilation

If the reference measurement is repeated and the door is opened wide, which corresponds to opening several windows, the curves shown in Figure 3 (grey dots) will result. This clearly shows that free ventilation is capable of reducing the virus load in the room in principle, but under these conditions it takes about 30 minutes until the virus load is halved. Furthermore, even small changes in the temperature differences cause the reduction to vary considerably, as the comparison of the two gray curves shows. The temperature difference between indoors and outdoors was less than 1.5 °C and the wind speed in front of the door was negligible. Therefore the low air exchange rate of 1.27 and 1.75 per hour is not surprising. The quantitative measurement results show beyond doubt that the method of free ventilation, as practiced in many schools, has its limits. In order to realize an air exchange rate of 6 per hour with the method of the free ventilation, at least 3.5 and/or 2.5 doors of the mentioned size would have to be permanently opened in the room with these temperature differences and wind conditions. It is questionable whether classrooms have such large window openings. It must also be taken into account that the outside and room temperatures are becoming more and more similar and therefore a large air exchange rate cannot be maintained permanently in practice. Due to the insufficient effectiveness of free ventilation, we do not consider safety concepts based on free ventilation alone to be safe.

We have already described as negligent the assertion of some HVAC system manufacturers that an air exchange rate of 1 – 2 should be sufficient to protect the children in the classroom from indirect infection, although air exchange rates of 12 – 15 are required in rooms with dangerous viruses. However, the results now illustrate very well how much the virus concentration in the room would actually decrease if this recommendation were followed. After one hour, the actual decrease of the virus load in this case is only 75%. It can hardly be said that this small decrease in the virus concentration can provide security against indirect infection. If one assumes that the HVAC systems are operated in winter with a very low proportion of outside air and the collection efficiency of the integrated filters of class F7 or F9 is significantly worse than that of a class H14 filter, it is clear that in this operating mode no security against indirect infection can be expected.

For comparison, the results with closed door and running air purifier clearly show the efficiency and advantage of the concept. With these devices, the aerosol particle concentration can be halved within 9 ½ minutes at an air exchange rate of 6 per hour set on the device. Therefore, high virus loads in the room can be quickly reduced with these units and kept at a very low level in the presence of virus sources. The filter performance is also completely independent of the season, window size and the willingness of children and teachers to open the windows.

The comparison of the function curves also clearly shows that it makes no significant difference whether the room is only furnished or whether people with bags, backpacks, laptops and jackets are also present in the room. Even the protection walls have no significant effect on the filter efficiency. Therefore, the objections that assume that the filter efficiency of the room air purifiers is significantly reduced in full rooms are wrong. The filter performance of the units can only be significantly reduced if the intake and discharge area of the unit is massively blocked [41]. But even a blocked room air cleaner is in most cases even more efficient than free ventilation.

From the curves of the aerosol particle concentration shown in Figure 3, the half-life and mean residence time can be calculated. The values for the different experiments are summarized in table 1. The comparison of the values shown in green shows that only after about 5 hours a reduction of the aerosol concentration by 50% in a room without ventilation is achieved. Free ventilation represents a significant improvement with 24 minutes, but even this value will not sufficiently reduce the risk of infection. The results show that with room air purifier the viral load is reduced by 50% within 9.6 minutes. A continuous filter operation will therefore reliably prevent an enrichment of the room air with infectious aerosol particles. Even several infected persons will probably not be able to create an infectious virus load in the room if the room air cleaner is operated with an air exchange rate of 6 per hour. Therefore, even under unfavorable conditions, the indirect risk of infection can be enormously reduced with these devices.

**Table 1:**
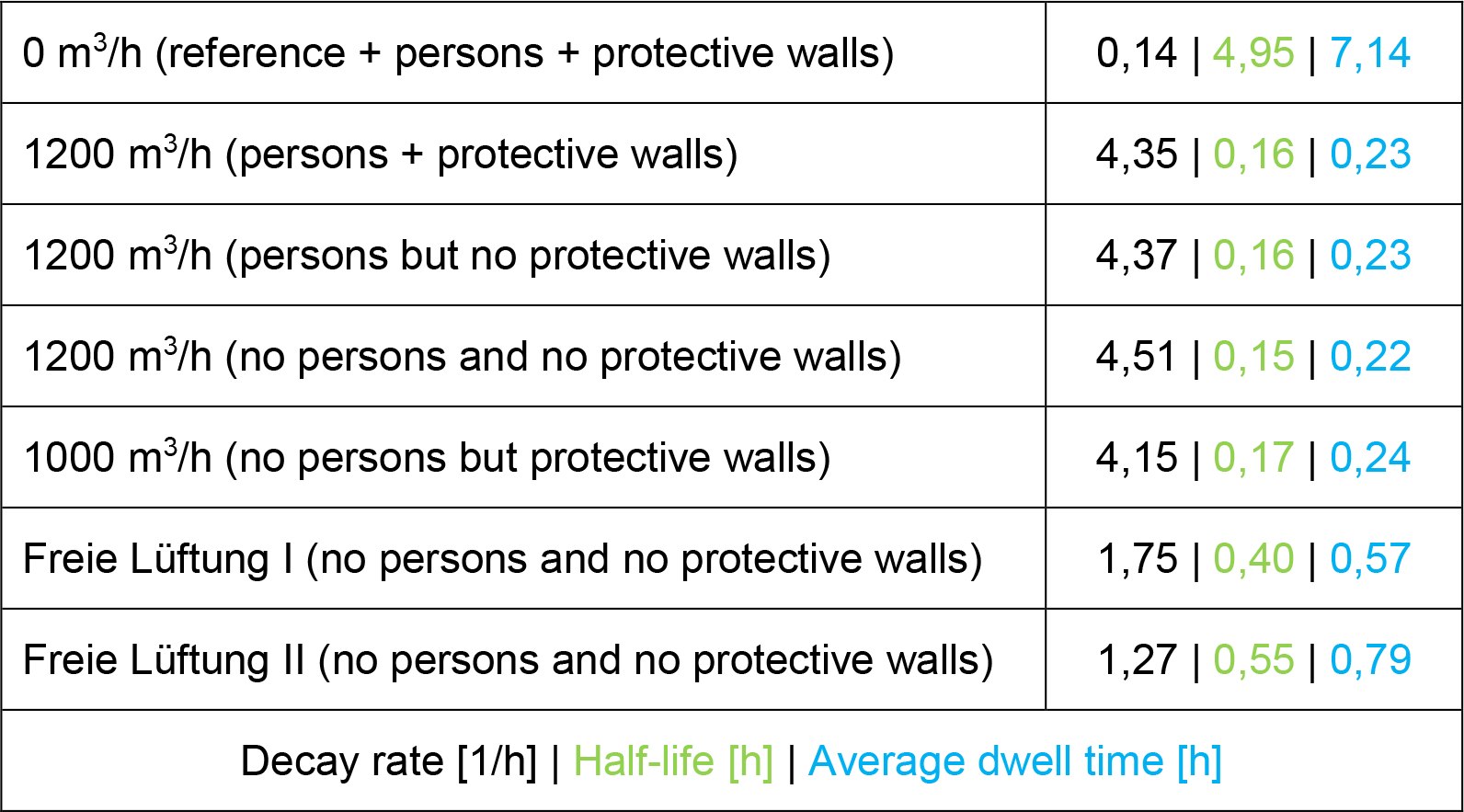
Decrease in aerosol concentration over time for different configurations. Decay rate (black), half-life (green) and mean residence time (blue).

If aerosol particles are continuously released in the room at the measuring position, it takes approx. 13 minutes, according to the information shown in blue, until they are streamed to the device and separated. If necessary, this value can be reduced by positioning the device more centrally in the room. Sometimes it is claimed that there is nothing better to do than to move the viruses through the room to the separation point with a room air purifier. This idea assumes that the viruses remain at the release site. But this is not the case, because the thermal and convective air movement in the room causes the viruses to be distributed homogeneously in the room after a short time. It is therefore much safer to guide the viruses to the filter and to separate them than to risk a distribution in the room.

### Experiments with local aerosol source

The experiments discussed so far make it possible to determine important quantities that are fundamentally important for qualifying the separation efficiency and the evaluation of the viral load in space. It could be argued that in reality an infected person in one place releases aerosol particles when breathing, speaking, singing, coughing and sneezing. A small preliminary test was conducted to determine whether the released aerosol particles could pass directly across the protection wall to the person sitting next to the seat. In the preliminary test, a table with a protection wall was positioned next to the measuring system, so that the measuring system could be used to analyse whether aerosol particles could reach the neighbouring seat while the room air purifier was running, if aerosol particles were continuously released at the neighbouring seat behind the partition wall. With the used aerosol generator it is not possible to release as few aerosol particles as a single person is able to do. The amount released could perhaps be realized by several dozen people. Thus the experiment analyses a case that is much more extreme than any real situation. The results shown in Figure 4 show that even with this enormously high release of aerosol particles, there seems to be no continuously increasing exposure to aerosol particles at the adjacent place. Instead, the values fluctuate around the initial concentration value. If the aerosol particles were to spread to the measurement location, a sharp increase in particle co-concentration would be expected. Therefore, it can be stated that these protective walls provide comprehensive protection against direct infection in classrooms or offices. The combination of an indoor air purifier and protection walls is therefore a very reasonable protection concept according to this study to achieve a very good protection against indirect and direct SARS-CoV-2 infection in classrooms and offices.

**Figure 4:**
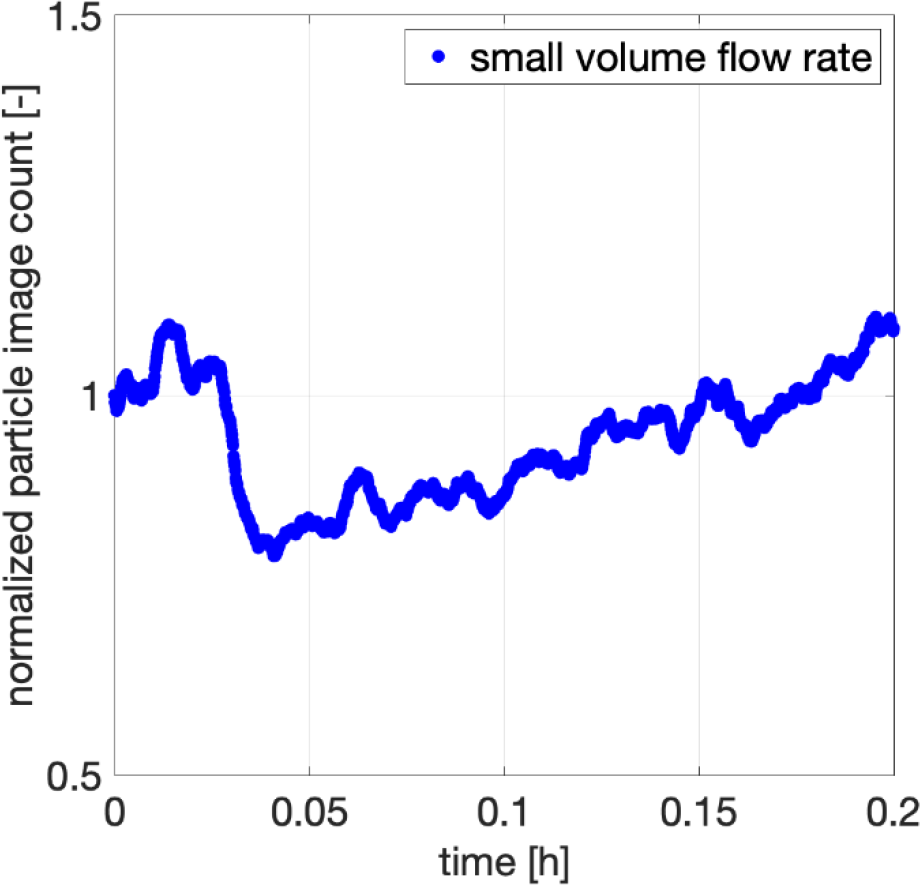
Aerosol particle concentration as a function of time with local release of aerosol particles behind the partition

## Summary and Conclusion

The main objective of this study was to compare different protection concepts that promise safe schooling during the SARS-CoV-2 pandemic and to determine whether a concept exists that is not only safe but also feasible, cost-effective and environmentally friendly. Due to the enormous importance of this issue for children and young people, but also for their parents and grandparents, we have collected missing data through scientific experiments to best secure our assessments. As a result the following can be determined and recommended:

A protection concept that relies solely on free ventilation is easy to implement, but it offers only a minimum of safety, since it does not provide any protection against direct infection. Infections are accepted cheaply with this concept. Infections cause enormous costs as soon as infected children and adolescents need medical care. But even if the parents cannot do their job because they have to take care of the children, e.g. if the class is in quarantine, this concept involves considerable follow-up costs. With this concept, the indirect risk of infection can be reduced during the warm season by means of free ventilation through permanently open windows or with HVAC systems that supply 100% outside air with an air exchange rate of 6 per hour. In winter, however, free ventilation is a pure waste of energy and therefore not an option, because infection prevention and climate protection must not be mutually exclusive. HVAC systems also offer no energy advantage in winter if they are operated with 100% outside air and six times the air exchange rate per hour. The protection concept is therefore unsafe, uncomfortable for children and young people due to the cold in the classroom, and economically and ecologically questionable. Consequently, the concept should not be implemented.

The protection concept, which relies on distances, cannot be implemented in reality because neither the room nor the teachers are available. The costs for the implementation would be massive and it would take a very long time to implement it, so that this concept can only be realized if the schooling takes place at home. However, this would not only be detrimental to the social development of the children and adolescents, but also enormously burdensome, since the parents would have to be very involved in the schooling process. It would also be economically inefficient, since parents would have to limit their professional activities and since the student-to-teacher ratio would be virtually 1 : 1 instead of 1 : 25. The concept is also unfair, since not all parents have the time, patience and qualifications to school the children and young people.

A protection concept that relies on high-quality particle-filtering respiratory masks (FFP2/3) offers a very high level of safety against infection. However, wearing them permanently has a negative effect on the health and well-being of the wearers. Therefore the concept is not really feasible. Furthermore, wearing respiratory masks makes neither economic nor ecological sense. The costs amount to 20000 € per year and class. In addition, a lot of waste is produced, which is ecologically unfavourable. However, it must be emphasized that these breathing masks must be worn as soon as the children leave their place and walk through class or school or go to their place in the morning or after the break!

Protection concepts in which the indirect risk of infection is realized by room air purifiers or disinfection devices have the advantage that the viruses in the room are separated or inactivated after a short time, provided (1.) the air exchange rate per hour is at least six times the room volume. (2.) 99.995% of the viruses are separated (with a class H14 filter) or inactivated (with UV-C or ionization) during a single pass through the unit and (3.) the unit is quiet so that it can be operated. Since the units locally remove or inactivate the viruses in the room air and do not waste heat energy, like free ventilation or air handling units, they can be considered as energy efficient. The technical implementation is simple and the devices are available in large quantities at short notice, since they are often manufactured in Europe. Therefore, their use would not only provide security against indirect infection, but also preserve jobs. The running costs are low due to the longevity of the filters (several years) and maintenance costs are not incurred for devices that work on a filter basis. Retrofitted installed HAVC systems however, are not an alternative. On the one hand, they are much more expensive than mobile room air cleaners and less energy efficient. On the other hand, their installation would require a long approval process, as two 400 mm core holes would have to be drilled into the outer walls of each classroom. Approval alone would not be possible in a year. Furthermore, fire protection and electrification would have to be clarified, as a simple power socket is not sufficient for these systems. Also there would not be enough craftsmen to install the systems at short notice. Mobile room air cleaners and disinfection devices do not have all these disadvantages. Therefore, government programs should be set up to enable schools to be equipped quickly with mobile air purifiers or disinfection devices.

Because the room air cleaners and disinfection devices minimize the indirect risk of infection, but do not provide protection against direct infection, additional protective measures are required. For the prevention of direct infections, mouth-and-nose covers, surgical masks or transparent face visors are suitable. Masks are often annoying and uncomfortable, they are unhygienic in the long run, are rarely worn properly and lead to waste. Face visors do not have these disadvantages and furthermore the facial expression is visible. However, they disturb the freedom of movement and the way of working. It is therefore advisable to position transparent protective screens with a surrounding edge between the seat neighbours. These are completely impermeable to aerosol particles and viruses, and if they are correctly dimensioned, it is also very unlikely that the exhaled air will rise, flow over the boundary and then sink down to the neighbouring seat. Partition walls therefore offer the best possible protection between adjacent people in the classroom. The table neighbours can get as close as they like through the barrier and can also talk and cough face-to-face without getting infected. Furthermore, the partitions do not disturb the students’ working methods, facial expressions are visible and the room is completely visible, provided that the protective walls are made of transparent Plexiglas. Thanks to the combination of the room air purifier or disinfection device and the protective wall, the children and adolescents can concentrate fully on the lessons and do not have to be afraid of infection, freeze with open windows or constantly think about the correct wearing of masks. In this way, a largely normal teaching operation can be realized, no matter how long the pandemic lasts.

The experiments carried out in this study confirm the very good effectiveness of the protective walls in preventing direct infections. Furthermore, we have shown experimentally that with powerful room air purifiers or disinfection devices a very efficient and fast filtering of the room air can be realized even if the room is full of people, bags, laptops and protective walls. From our point of view this protection concept offers a high degree of safety, is easy to realize, relatively inexpensive if the costs of the other concepts are seriously considered, and ecologically sustainable. Therefore, based on this analysis and experiments, we recommend a step-by-step implementation of the concept, taking into account our recommendations.

If this concept is implemented, the question remains what to do with the equipment after the pandemic. From our point of view, it can be assumed that these devices will continue to be used after the current pandemic to combat other infectious diseases, but also to permanently remove fine dust and pollen from the air in order to protect allergy sufferers and people’s health. In the long term, the knowledge gained here may also be reflected in HVAC systems, so that in future these systems will take over the task of mobile room air cleaners and disinfection devices. But this is a very long-term process.

## Note

The investigations were financially supported by the company TROTEC GmbH, Heinsberg. The room air cleaner TAC V+ was provided by TROTEC for the investigations. The investigations were carried out in accordance with good scientific practice. The support provided by TROTEC has no effect on the results presented.

## Data Availability

The data is stored on an internal server and can be provided on request.

## Notes

### Competing Interest Statement

The investigations were financially supported by the company TROTEC GmbH, Germany. The room air cleaner TAC V+ was provided by TROTEC GmbH for the investigations. C.J. Kähler received personal allowances as part of his payment as a professor.

### Author Declarations

In the study, no experiments were carried out on humans or living creatures, so this point does not apply or is closed.

